# Female genital schistosomiasis and reproductive tract infections. A cross-sectional study in rural adolescents in South Africa

**DOI:** 10.1101/19009233

**Authors:** Jilna Dilip Shukla, Elisabeth Kleppa, Sigve Holmen, Patricia D. Ndhlovu, Andile Mtshali, Motshedisi Sebitloane, Birgitte Jyding Vennervald, Svein Gunnar Gundersen, Myra Taylor, Eyrun Floerecke Kjetland

## Abstract

**Background and objectives:** The aim of the current study was to establish the relative prevalences of Female Genital Schistosomiasis (FGS) and sexually transmitted infections (STIs). We hypothesised that due to the use of syndromic management for STIs it is possible that FGS is being misdiagnosed and mismanaged as an STI. We therefore wanted to examine the relationship between FGS and the individual STIs in schistosomiasis endemic areas.

**Methods:** Between 2011 and 2013, a cross-sectional study was performed in 32 randomly selected secondary schools in rural KwaZulu-Natal, South Africa, where each school had at least 300 pupils. In a research clinic, FGS diagnosis, STI testing, and face-to-face interviews were performed in sexually active, young women aged 16 – 22 years.

**Results:** FGS was the second most prevalent current genital infection (23%). There were significantly more women who had presented FGS among those who had detectable urinary schistosomiasis (35%), compared to those without (19%, p< 0.001). In the FGS positive group 35% were positive for HPV infection, compared to 24 % in the FGS negative group (p=0.010). In the FGS positive group 37% were sero-positive for HSV infection, compared to 30% in the FGS negative group (p=0.079). There were significantly fewer chlamydia infections amongst women with FGS (20%, p=0.018) compared with those who did not have FGS (28%).

**Conclusions:** FGS was the second most common genital infection after HSV but the two were not significantly associated. HPV infection was significantly associated with FGS. Surprisingly Chlamydia infection were negatively associated with FGS. The results show the importance of the inclusion of FGS in the management protocols for genital infections in areas endemic for urinary schistosomiasis, and highlight the importance for more research on suitable differential diagnostic tools and disease management.

**Key messages box:**

- FGS was the second most common genital infection in this rural population after HSV.
- FGS was positively associated with HPV.
- FGS was negatively associated with genital chlamydia infections.
- FGS should be included in the syndromic management of genital infections.

## Introduction

Female genital schistosomiasis (FGS) was first described in 1899 in Egypt but was largely only reported as incidental findings till the 1990s.^1^ It has been found to be almost as common as urinary schistosomiasis within the female populations of endemic areas.^2–4^ Furthermore, in some schistosomiasis endemic communities it may be the most common gynaecological morbidity.^4^ Despite its prevalence, World Health Organisation (WHO) data for 2017 shows that only 46.3% of the 220.8 million people requiring treatment for schistosomiasis have been reached.^5^

Genital schistosomiasis is a poverty-related neglected tropical disease caused by the *Schistosoma haematobium* parasite, which is transmitted by contact with infested water.^4^ It is known to be endemic in areas of sub-Saharan Africa where lakes, rivers and dams are used for domestic and recreational purposes.^6^ FGS may cause blood stained and foul smelling discharge, dyspareunia, contact bleeding, burning sensations in the genitals, decreased fertility and spontaneous abortions.^7–10^ The *S. haematobium* parasites inhabit blood vessels surrounding the female genital tract and the urinary bladder.^4^ There they lay eggs, which then migrate towards the adjacent organs, creating the afore mentioned morbidities.^4^ The morbidity seen in FGS is attributed to chronic inflammation in the cervical and vaginal mucosa and lesions involve both dead and viable parasite eggs.^11^

FGS is a differential diagnosis to some STIs, causing vaginal discharge and ulcers.^12^ Furthermore, it has also been hypothesised that FGS may make females susceptible to Human papillomavirus infection and other sexually transmitted diseases.^13,14^

In this study, we aim to explore the relationship between FGS and reproductive tract infections in an adolescent female population and establish the relative prevalences of these infections in a rural *S. haematobium* endemic area.

## Methods

### Study Design

It has been shown that the schistosomiasis problem is in the lower altitudes KwaZulu-Natal Province of South Africa.^15^ Therefore, secondary schools below the altitude of 400 meters were invited to take part in the study. Nested in a prospective study on the prevention of FGS, the study included schools with a urinary schistosomiasis prevalence above 20%.^16,17^ For practical purposes and due to financial constraints, we only invited schools with three hundred or more pupils. Schools were excluded if they had been classified as urban (and therefore less likely to be using infested water sources) by the Department of Education. From 2011 to 2013 we included sexually active school attending adolescent women aged 16-22 years, usually in grades 10-12, who consented to a gynaecological examination, and were willing to be tested for STIs.^17^

### Clinical Examination and Questionnaire

Trained female staff interviewed study participants in isiZulu (the local language) using a questionnaire that covered demographic information, current reproductive health complaints, obstetric history, sexual behaviour, water contact and their approaches to the health services for genital symptoms. Participants then underwent a gynaecological examination performed by female doctors trained in recognising FGS. Clinical signs of FGS were documented by photo-colposcopy (Olympus OCS 500 colposcope with a mounted Olympus E 420 10 megapixel (Mpx) single lens reflex (SLR) device, or a Leisegang colposcope with a Canon EOS 40D 10 Mpx SLR).^18^ Grainy sandy patches were defined as lesions with grains measuring approximately 0.05 ⨯ 0.2 mm and shaped like minuscule rice grains, deeply or superficially situated within the mucosa.^12,19^ Homogenous yellow patches were defined as yellow areas with no distinct grains. Rubbery papules were defined as beige to yellow papules with a firm (rubbery) protrusion into the lumen. All the FGS lesions may be accompanied by characteristic abnormal blood vessels.

### Laboratory testing

For the diagnosis of urinary schistosomiasis, a urine sample was collected between 10 a.m. and 2 p.m. which is the peak daily egg excretion time.^20^ Urine was centrifuged, the sediment deposited on two slides and read by two independent laboratory technicians.

Biopsies were not taken because the post-biopsy lesion may pose an unnecessary raised risk of HIV transmission in this HIV endemic area.^21^ Urine was considered schistosomiasis positive if at least one *S. haematobium* ovum was seen under the microscopy. Samples of cervicovaginal lavage (CVL) was collected by spraying 10 mL of saline on the cervical surface and withdrawing it back into the syringe. *Herpes simplex virus* type 2 (HSV-2) antibodies were detected in serum using ELISA Ridascreen HSV-2 IgG, (Davies Diagnostics, Randburg South Africa) and in ELISA HerpeSelect®, IgG assay, (Focus Diagnostics, Germany). *Herpes simplex virus* type 2 PCR was only done in a subsample of participants due to financial and practical constraints. Likewise, Human Papillomavirus (HPV) was done in a sub-sample, and detected by GP5+/6+ HPV PCR followed by an enzyme immunoassay method using a cocktail mix HPV probes (Whitehead Scientific South Africa).^22^ Cervical swabs were analysed using ProbeTec CT/GC and AC strand displacement PCR assay for *Chlamydia trachomatis* and *Neisseria gonorrhoea* (Becton Dickinson, Microbiology systems, United States).^17,18^ In-house PCR was used for the screening of *Trichomonas vaginalis* and HSV, in the Laboratory of Infection, Prevention and Control at the University of KwaZulu-Natal, Durban, South Africa.^23^ Syphilis was detected in thawed serum samples using Macro Vue^™^ test 110/112 (BD, Becton Dickinson Microbiology systems, United States) for a rapid plasma reagin test (RPR) and Immutrep for *Treponema pallidum* haemagglutination assay (Omega diagnostics group PLC, Alva, Scotland, UK). Nugent’s scoring criteria was used for the diagnosis of bacterial vaginosis. Candidiasis was diagnosed using the Gram staining technique under light microscopy, and severity scored (one to five) in accordance to the number of spores seen.^24^

### Ethics

The study was approved by the Biomedical Research Ethics Administration (BREC), University of KwaZulu-Natal, South Africa (Ref BF09/07) and the Department of Health, Pietermaritzburg, South Africa (Ref HRKM010-08). Ethical clearance was also granted from the regional committee of Medical and Health Research Ethics (REC) South Eastern Norway (Ref 469-07066a1.2007.535) and renewed in 2011. Specimens were collected after obtaining a written informed consent and each young woman underwent recruitment procedures as described previously.^17^ For participants who were 16-17 years of age, the ethics committee granted permission for independent minor consent without parental consent. Permissions were also granted by the Departments of Health and Education, KwaZulu-Natal.

### Statistical Analysis

In order to be able to detect a 10% difference in an STI’s prevalence between the FGS positive and negative groups with a confidence level of 0.05 and power of 0.8, sample size calculations indicated that we would need 200 FGS positive patients and 600 FGS negative patients. Chi-Square or Fisher’s exact tests were used to evaluate the hypothesis. Age was included in the multivariable logistic regression regardless of significance level. Other variables were included if the association between the sandy patches and the STIs had a p-value lower than 0.2. Comparison of mean age between the FGS positive and negative groups was done using the Student’s T-test. Analyses were performed using IBM SPSS Statistics Version 25 (Armonk, NY, US).

## Results

A total of 933 participants from 32 schools were willing to be tested for STIs (Figure 1). One or more STIs were found in 819/930 (88.1%) of the participants and FGS was found in 210/933 (22.5%). More than half of these rural secondary school going young women had been pregnant (Table 1). The mean age was 18.7 years (Standard Deviation (SD) 1.6) indicating that approximately half of them had missed a year or more of schooling. Most women had been exposed to schistosomiasis risky water at some point in their lives 881/930 (94.7%). Urinary *S. haematobium* ova excretion was found in 256/840 (30.5%). In the positive cases the geometric mean intensity urinary *S. haematobium* was 57.9 eggs per 10 millilitres. There was no difference in mean age of those who had FGS compared to those who did not. Evidence of a prior HSV infection (serology) was found in 287/900 (31.8%) of the participants, and the prevalences of current chlamydia, HPV and trichomoniasis were 218/827 (26.4%), 149/551 (27.0%) and 163/839 (19.4%) respectively.

**Table 1.** The main characteristics of participants

| **Variable** | **Number of participants n (%)** |
| --- | --- |
| Urinary schistosomiasis<sup>a</sup> | 256 ( 30.5) |
| FGS manifestation<sup>12,19</sup> |  |
| - Any sandy patches of cervix or vaginal wall | 210 (22.5) |
| Superficial grainy sandy patches | 84 (9.0) |
| Deep grainy sandy patches | 51 (5.5) |
| Homogenous yellow sandy patches | 109 (11.7) |
| - Rubbery papules | 0 (0) |
| - Abnormal blood vessels | 277 (29.7) |
| Lifetime risk water contact.<sup>b</sup> | 884 (94.7) |
| **Religions and language** |  |
| Christian | 712 (76.3) |
| Shembe<sup>c</sup> | 184 (19.7) |
| Other/ no religious affiliation | 20 (4) |
| IsiZulu language spoken at home | 923 (98.9) |
| **Highest education level amongst family members (not participants)** |  |
| Tertiary<sup>d</sup> education | 97 (10.4) |
| Secondary<sup>e</sup> education | 827 (88.6) |
| Primary<sup>f</sup> school/no education | 2 (0.2) |
| **Obstetric history** |  |
| Previously pregnant | 478 (51) |
| Parity (Median (IQR)) | 1 (1-3) |
| <sup>a</sup>*S. haematobium* ova found in a single urine sample. <sup>b</sup>River, dam, pond or stream. <sup>c</sup>Local popular religion. <sup>d</sup>University level. <sup>e</sup>8-12 years of school. <sup>f</sup>1-7 years of school. |  |

**Figure 1.**
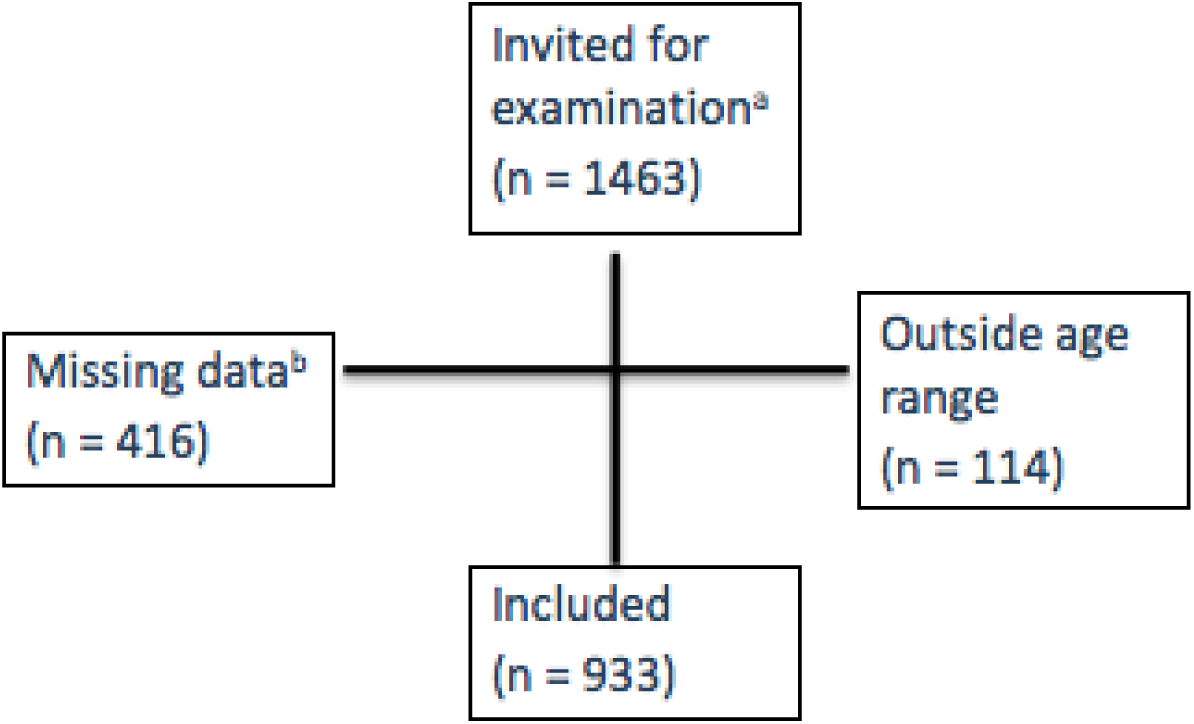
Inclusion criteria. ^a^Female pupils in schools with more than 20% urinary schistosomiasis, grades 10-12, sexually active. ^b^Did not undergo gynaecological examination (n=416), did not submit a questionnaire (n=42), did not submit urine (n=295) and 301 had overlapping reasons for being excluded.

HPV testing was done in a subsample of 551 patients (55%) and was positively associated with FGS. When HPV was analysed against the sub-types of sandy patches, it was positively associated with superficial grainy sandy patches (Age Adj. OR 1.85, 95% CI (1.05 – 3.245) p = 0.03) and homogenous sandy patches, (Adj. OR 2.43, 95% CI (1.49 – 4.0) p < 0.001). HPV was however, not associated with deep grainy patches (Age Adj. OR 0.70, 95% CI (0.30 – 1.7) p = 0.42), but the sample size was small (51 cases).

There was significantly less *Chlamydia trachomatis* in the FGS positive group than in the FGS negative, after controlling for age, HSV serology and trichomoniasis. Chlamydia was also negatively associated with superficial grainy sandy patches (Age Adj. OR 0.44, 95% CI (0.26 - 0.75) p = 0.002) and deep grainy patches (Adj. OR 0.30, 95% CI (0.12 - 0.77) p = 0.01). Homogenous sandy patches showed the same tendency, (Adj. OR 0.71, 95% CI (0.42 -1.19) p = 0.19). Similarly, the chlamydia prevalence tended to be lower in the urinary schistosomiasis positive cases than in urinary schistosomiasis negative, although this difference was not significant (Adj. OR 0.70, 95% CI (0.45 - 1.02) p = 0.06). Half, 109/218 (50.0%) of the *Chlamydia trachomatis* cases were clustered in 8/32 (25.0%) schools. After stratification, taking out the chlamydia high-endemic schools, FGS remained negatively associated with chlamydia, after adjusting for age, trichomoniasis and HSV serology, (Adjusted Odds Ratio (Adj. OR) 0.41, 95% CI (0.22 – 0.79), p = 0.007) but not in the eight schools, each of which had more than 35% chlamydia. Of the study participants 12.6% (110/876) had been treated for a sexually transmitted infection previously, there was a significant association with having HSV (OR 1.5, 95% CI 1.2-1.9, p=0.001) but no association was found with FGS (p=0.97), chlamydia (p=0.33), dual infection with FGS and chlamydia (0.45) or other STIs.

*Trichomonas vaginalis* was associated with the clinical finding of “homogenous sandy patches” (Age Adj. OR 1.74, 95% CI (1.10 - 2.75) p= 0.019). No other STIs were associated with grainy sandy patches (data not shown). Positive HSV serology and current trichomoniasis were higher in the FGS positive group than in the FGS negative but the differences were not significant. The mean bacterial vaginosis scores were the same in FGS positive and negative populations. Only, 29/901 (3.2%) cases were positive for *Treponema pallidum* and there was no significant difference between the groups (p =0.49).

HSV PCR (indication current shedding) was done in a subsample and only 12/351 individuals were positive. There was no significant difference in HSV shedding in FGS positive participants compared to negative cases (OR 0.95, 95% CI (0.33 - 2.76), p=0.92).

As expected, FGS was positively associated with urinary schistosomiasis (OR 2.32, 95% CI (1.77- 3.02), p < 0.001). FGS was found in 89/256 (34.8%) of those who were positive for urinary schistosomiasis but also in 110/584 (18.8%) of the participants where urinary schistosomiasis was not detected (only one urine sample, for each participant examined). Amongst the five participants who denied any waterbody contact, none were diagnosed with urinary schistosomiasis although sandy patches were found in one case. Urinary schistosomiasis was not associated with any of the other reproductive tract infections.

As further exploration, we included HPV in the multivariable analysis of Table 2. HPV remained significantly associated with FGS (Adj. OR 1.71, 95% CI (1.11 – 2.38), p = 0.016). However, there was less chlamydia in the FGS positive than in the FGS negative but the difference was not statistically significant (Adj. OR 0.67, 95% CI (0.42 – 1.07) p = 0.096).

**Table 2:**
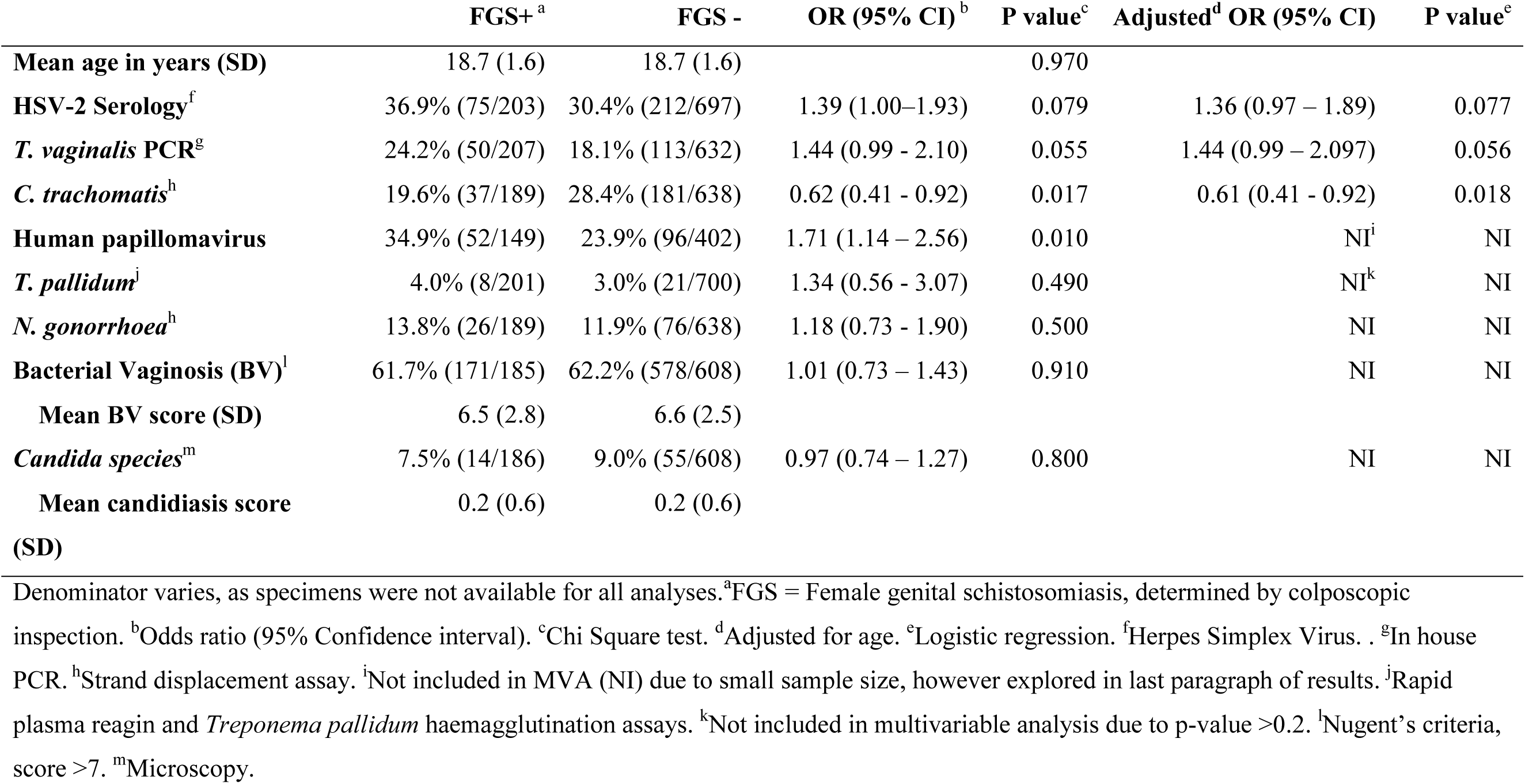
The association between sexually transmitted infections and Female Genital Schistosomiasis in young women

## Discussion

Female Genital Schistosomiasis was almost as common as chlamydia, with both infections affecting more than one fifth of this young rural population. Unexpectedly, the FGS positive had less concurrent chlamydia infection, compared to those without FGS. This has not been reported previously. As published previously, the study shows that FGS seems to be a risk factor for HPV.

FGS manifestations cause a chronic state of intravaginal inflammation which could hypothetically aid the acquisition of HPV and other STIs.^11,25^ The *S. haematobium* transmission and manifestations as FGS are likely to have occurred in early childhood and therefore prior to sexually transmitted infections.^10,26^ Previous studies indicate that there are intravaginal damages already from a young age. If the vaginal discharge has been constant since childhood, it could be argued that it would be interpreted as normal by the patients.^10,26^ However, one study found that adults with FGS reported having abnormal discharge.^27^ This may indicate that they have experienced changes in discharge, possibly because the bleeding and ulcerous surfaces of FGS are super-infected by other agents such as bacteria, as has been found for *S. haematobium* of the bladder in mouse models.^28^

There was an overall trend of higher prevalence of sexually transmitted infections in the FGS positive participants. However, chlamydia was the exception. It could be hypothesised that FGS somehow protects women from chlamydia. However, could be hypothesised that young women with FGS and chlamydia sought STI treatment provided freely by the local clinics, possibly due to intensified symptoms with dual infection.^29^ This could not be confirmed in our study.

In order to detect small differences between the groups, we would have had to have a larger sample size. Almost all participants in this population had water contact at some point in their lives. Due to logistic and resource constraints, only one urine sample was collected per participant. Multiple samples would likely have yielded a higher prevalence of urinary schistosomiasis due to day to day variation in egg excretion.^30^ This was a limitation in this study. Furthermore, FGS diagnosis is dependent on the subjective inspection of all intra-vaginal surfaces, including the fornices and posterior and anterior vaginal walls.^12,19^ Therefore we may have also have missed some FGS positive cases. As is often the case in schistosomiasis research, this might have obscured differences between the FGS positive and FGS negative groups.^3^

In rural South Africa, abnormal discharge and genital ulcers are currently managed syndromically as STIs, without laboratory testing.^31^ The symptoms of FGS overlap with symptoms of the various STIs causing discharge, a burning sensation in the genitals and ulcers.^10,27^ However, in countries where FGS is endemic, laboratory testing for STIs and examinations are often not performed. The current syndromic management for vaginal discharge entails treatment for chlamydia, gonorrhoea and trichomoniasis, but not for FGS.^31^ Therefore, patients may be inadvertently over-treated for the STIs and undertreated/not treated for FGS.

Examinations for FGS rely on clinical expertise, and are not yet performed in rural healthcare.^12,25^ The risk of misdiagnosis of FGS as an STI warrants rewriting the syndromic management protocols and algorithms for genital tract disease, especially in endemic areas. Furthermore, symptoms of STIs and FGS are also found in children with *Schistosoma haematobium*.^10^ Yet health professionals are not taught about FGS, and FGS is not on the list of differential diagnoses for genital ulcers and discharge. Therefore changes in the health professionals’ curricula and the protocols for management of reproductive tract diseases in females are necessary.^21,32^ Anti-schistosomal treatment should be added to the treatment protocols and FGS should be considered amongst the differential diagnoses.

Preventive treatment for *S. haematobium* with praziquantel is not expensive and readily available and may provide a reduction in FGS occurrence. Further research is needed to understand the physiological mechanisms behind the sandy patches and to explore if praziquantel treatment can reduce the incidence of HPV and subsequent cervical cancers.

## Data Availability

Data will be available, however it is still undergoing publication and facts are being used by other authors.

## Acknowledgements

We wish to thank the Departments of Education and Health who saw the importance of this work. Teachers, parents and pupils in schools in KwaZulu-Natal Province are thanked for their kind assistance. Thanks also to staff members of the BRIGHT research team who treated young women with kindness and respect, drove long distances, worked late hours, lived under harsh conditions and bent over backwards to accommodate many unexpected events, in alphabetical order, B. Ndlovu, E. Kwela, G. Hlengwa, J. Hargreaves, L. Radebe, M. Majiya, N. Mbiza, N. Mpofana, N. Msomi, N. Mkhabela, P. Cele, R. Manyaira, S. Nzimande, S. Sibiya, S. Gagai, T. Mqadi, T. Mkhize, V. Duma, Z. Chiliza, Z. Ndovela.

